# Implementation of Lot Quality Assurance Sampling Using Mobile Data Collection Tools to Assess Vaccination Coverage After Synchronized Polio Vaccination in the Province of Camiguin, Philippines

**DOI:** 10.1101/2020.03.17.20033571

**Authors:** Ian Christian A. Gonzales

## Abstract

**Objectives:** The study assessed immunization coverage after one round of synchronized Polio vaccination in Camiguin, Philippines. It included classifying the coverage level, identifying the level of awareness and source of information for the campaign, describing the reasons for non-vaccination, and pilot a mobile data collection platform.

**Methods:** Lot Quality Assurance Sampling (LQAS) is a household survey using a multi-stage clustered technique. Sixty respondents per municipality, divided into 6 clusters, with one barangay considered as one cluster. Barangays were selected with probability proportional to size. Households were taken by systematic sampling. One child was randomly selected if there were multiple children in a household. Data collection was done using KoBo Toolbox.

**Results:** The municipalities of Mahinog and Sagay had two unvaccinated children each. Guinsiliban and Mambajao only had one unvaccinated child each. Catarman did not have any unvaccinated child. The reasons for non-vaccination were the lack of means of validation, fear of side effects, and the absence of a caregiver at the time of vaccination. The most common sources of information were health workers and television.

**Discussion:** Coverage for all five municipalities of the province passed the decision value. The main reason for non-vaccination was the lack of means of validation, which emphasizes the need for high quality finger marking and the provision of vaccination cards. Only Mahinog did not pass the threshold for campaign awareness. LQAS is useful for validating areas with concerns on the set target population and administrative coverage. Mobile data collection through KoBo ToolBox is a useful method for field use. It is easily adaptable, user-friendly, and allows for immediate data validation and analysis.

## INTRODUCTION

The last known case of wild Poliovirus in the Philippines was in 1993.^1^ The Philippines has been Polio-free since October, 2000. However, environmental samples taken from Tondo, Manila last July and August, 2019, were found positive for Vaccine-Derived Poliovirus Type 1 (VDPV1). On September, 2019, VDPV1 and Vaccine-Derived Poliovirus Type 2 (VDPV2) were again detected in Tondo, Manila and VDPV2 was also detected in an environmental sample from Davao City. In addition, VDPV2 was confirmed at Lanao del Sur in a three-year-old child with Acute Flaccid Paralysis (AFP) symptoms. The isolated VDPV2 was found to be genetically linked to the 2 confirmed VDPV2 environmental samples. As a result, VDPV2 was classified as circulating (cVDPV2). The Department of Health (DOH) confirmed the re-emergence of polio in the Philippines and declared a national polio outbreak, in line with International Health Regulations (IHR).^2^ As of 12 December 2019, there were nine confirmed cases of Poliovirus. In response,synchronized Polio vaccination was done for Mindanao and the National Capital Region (NCR).^3^

Round one was recently conducted last 25 November to 13 December, 2019. For the Northern Mindanao region, 554,239 out of 584,076 targeted children aged 0-59 were vaccinated with an administrative coverage of 95%. The target population was computed based on projected population.^4^

Camiguin is an island province in the Philippines with a population of 88,478 as of the 2015 census. It has five municipalities: Catarman, Guinsiliban, Mahinog, Mambajao (Capital), and Sagay.^5^ For the campaign, the target population of 0-59 months old children was 10,203 and local health workers were able to vaccinate 8,409 children, for an administrative coverage of 82%.^4^ Local health workers cited the high projected population over the actual population which reduced their administrative coverage.

In the conduct of synchronized Polio vaccinations, a quality assurance tool is important to identify and target low-coverage areas, recognizing the large investment in manpower and financial resources to conduct a massive campaign. The Lot Quality Assurance Sampling (LQAS) was developed to quickly assess the performance of vaccination campaigns at the local level. LQAS provides classification of areas in terms of pre-defined vaccination coverage levels. However, it does not provide a point estimate of vaccination coverage.^6^ Other methodologies used to assess coverage is the Rapid Coverage Assessment (RCA), also called Independent Monitoring (IM), and Expanded Program for Immunization (EPI) Cluster Surveys. The table below compares the advantages and disadvantages of each methodology:

**Table 1.**
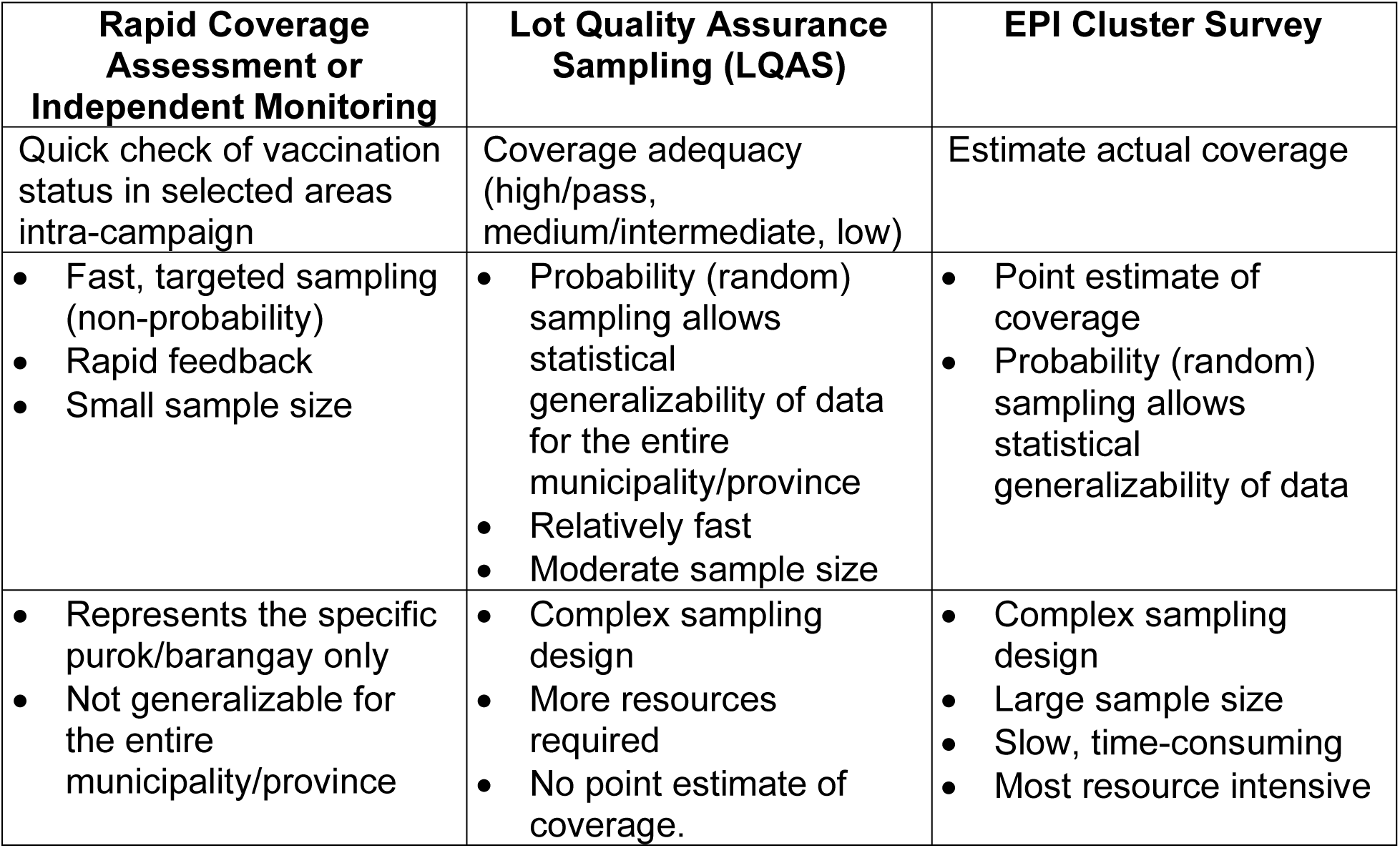
Comparison of vaccination coverage assessment methodologies.

Monitoring and evaluation of outbreak response requires relevant data in a quick time. Study designs employing larger sample sizes may not be appropriate except for end-term evaluation while non-probability sampling strategies are not preferred for lack of their representativeness.^7^ The LQAS methodology strikes the best balance between ease of field implementation and statistically-reliable results to track trends over time.^8^ Recent outbreak response immunizations for Measles and Polio in the Philippines were only assessed through RCA/IM. Upon review of literature, there are no publications from the Philippines where post-immunization coverage was assessed using LQAS.

The lot is the area of intervention. If a lot is rejected by the LQAS rule then mop-up vaccination will be implemented at this level. Hence, it is generally based on the administrative level in which the action can be taken.^5^ In the Philippines, this is an area where a local health officer has supervision, such as a city or municipality. Clusters can be defined as the localities with clear administrative boundaries present in the lot, such as a barangay.

Mobile data collection is advantageous over paper-based forms because of the rapidity of deployment, reduced errors in encoding and consolidation, and instant availability of the data for analysis. Those features are important especially in emergency settings such as outbreak response. KoBo Toolbox is an adaptation of the Open Data Kit (ODK), which is an open-source platform for electronic data collection using mobile devices. This system uses a mobile application for Android devices or the Safari web browser for iOS devices. It allows for both online and offline data collection which is advantageous in areas with poor cellular signal.^9^

The general objective of the study was to assess the quality of the Round 1 synchronized Polio vaccination campaign in the province of Camiguin. Specifically, this study aims to:

1. Classify the coverage level of each municipality.
2. Identify the level of awareness and source of information regarding the campaign.
3. Describe the reasons for non-vaccination if unvaccinated children are found.
4. Pilot KoBo Toolbox as an mobile data collection platform for the survey.

The data collected from the LQAS will help improve the microplans of municipalities in Camiguin for subsequent vaccination campaigns, specifically in reaching unvaccinated children and improving risk communication to increase campaign awareness and reduce non-vaccination. The application of mobile data collection tools will allow more efficient monitoring and evaluation in future outbreak response.

## METHODS

The LQAS was a household survey with sampling methodology based on guidelines from World Health Organization (WHO) and Global Polio Eradication Initiative (GPEI).^6,10^ Multi-stage clustered sampling was used to improve the practicality and rapidity of the study. The municipality was identified as the lot, and all five municipalities of Camiguin were included in the study. A sample size of sixty (n=60) children for each municipality were taken. The sample for each municipality was divided into 6 clusters, with one barangay considered as one cluster. Barangays were randomly selected with probability proportional to size (PPS). Within each barangay, a purok was selected using a randomizer application and ten eligible households were selected based on systematic sampling with a random starting point. A household is considered eligible when at least one child aged 0-59 months was present at the time of the visit. When multiple children were present within a household, one child was selected for the survey using a randomizer application.

Vaccination status was validated by either of three methods: finger marking, vaccination card, or master list. Verbal history without other means of verification was not considered. Data collection was done using KoBoCollect application for Android devices or Enketo using Safari browser for iOS devices. The server and electronic form were created through KoBo Toolbox. The survey was conducted from 17-19 December, 2019. The study was ethically reviewed and approved by the Management Committee of the Center for Health Development – Northern Mindanao, Department of Health. The study was carefully explained and the caregiver’s consent was secured by enumerators prior to the actual data collection.

## RESULTS

A total of 324 children were included in the study. Majority of the children were males (n=179). The median age is 30 months old (range = 1 to 58 months).

**Table 2.** Coverage and awareness classification per municipality (n=5).

| Municipality | Number of Unvaccinated | Coverage Classification | Number of Unaware Caregivers | Awareness Classification |
| --- | --- | --- | --- | --- |
| Catarman | 0 | PASS | 1 | PASS |
| Guinsiliban | 1 | PASS | 1 | PASS |
| Mahinog | 2 | PASS | 7 | FAIL |
| Mambajao | 1 | PASS | 1 | PASS |
| Sagay | 2 | PASS | 1 | PASS |

In the entire province, there were six unvaccinated children found.

**Table 3.** Unvaccinated children in the province of Camiguin, n=6.

| Age in months | Sex | Municipality | Reason for non-vaccination |
| --- | --- | --- | --- |
| 24 | Male | Guinsiliban | Unaware of the campaign<br>Parent/guardian was not present at the time |
| 18 | Male | Mahinog | Vaccine team did not visit |
| 55 | Male | Mahinog | Child was not from Camiguin, no finger marking |
| 40 | Male | Mambajao | Vaccinated as claimed but no finger marking and not in master list |
| 41 | Male | Sagay | Child was not from Camiguin, no finger marking |
| 52 | Male | Sagay | Fear of side effects by grandparents but mother will allow vaccination during round 2. |

The most common means of validation was through finger markings (n=169). For children who no longer had finger markings, their status was validated through a master list provided by the LGU (n=136). A notable good practice for the municipality of Sagay was to put a sticker on the door which indicated the number of eligible children, vaccinated children, and date of vaccination.

The two most common source of information were health workers and television.

**Table 4.** Source of synchronized polio vaccination campaign information, multiple responses per respondent, n=313.

| Source of Information | Number of Respondents |
| --- | --- |
| Health workers | 295 |
| TV | 108 |
| Barangay officials | 36 |
| Radio | 31 |
| Streamer/tarpaulin | 26 |
| Relatives/neighbors | 26 |
| Social media | 10 |
| Flyers | 3 |

## DISCUSSION

### Vaccination Coverage

The approach recommended by the Global Polio Eradication Initiative (GPEI) to assess the quality of Oral Polio Vaccination (OPV) coverage is based on a “band approach” that allows us to classify lots in three bands (High/Pass, Medium/Intermediate, and Low coverage) using two decision values (d). The recommended interpretation framework allows classification of the lots, with thresholds based on two levels of coverage (90% and 80%) and two decision values (3 and 8) using a sample of 60.^6^

**Figure 1.**
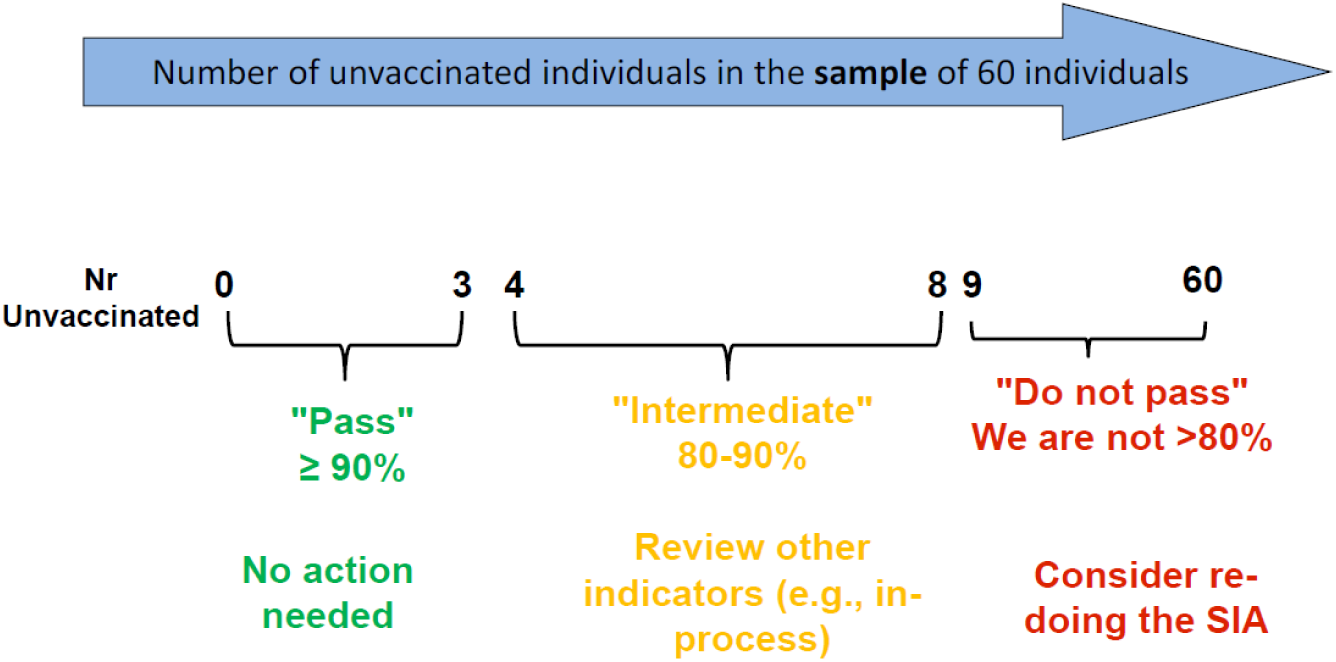
Interpretation framework per lot.^6^.

Each municipality had two or less unvaccinated children. This validated the feedback of local health workers that there are very few unvaccinated children to be found. Given these results, all municipalities of Camiguin were classified as PASS with a level of coverage ≥90%. This was in contrast to the province’s administrative coverage of only 82%. Because the projected target population serves as a denominator for computing administrative coverage, areas with a lower actual population will have lower administrative coverage. Although RCA/IM done in Camiguin showed an average coverage of 99%, results are usually biased upward.^8^ Inherent with the non-probability sampling of RCA/IM, results should not be generalized for an entire municipality or province. LQAS is therefore an indispensable tool for validating these areas with statistical reliability, with a better capacity to differentiate areas with low vs. high quality compared to the RCA/IM methodology.^8^

### Campaign Awareness

In parallel to vaccination activities, intensified risk communication and social mobilization take place prior to and during the campaign. The decision rule to identify whether a lot has adequate campaign awareness was also based on GPEI recommendations:^6^

- 0-3 not aware: awareness may be reaching 90% → PASS
- 4 or more not aware: not confident that awareness is above 90%→ FAIL

It is notable that most of the caregivers obtained their campaign information from health workers, which means that there was adequate on-the-ground risk communication during the pre-implementation phase. It is only in the municipality of Mahinog where the number of unaware caregivers exceeded the threshold. Intensification of risk communication activities for the municipality of Mahinog is therefore recommended. There are still avenues which are underutilized, such as dissemination through barangay officials who may have more influence over their constituents and/or the addition of more tarpaulins/streamers to increase campaign visibility.

### Reasons for Non-Vaccination

Of the six unvaccinated children found in the province, two claimed that they were vaccinated but were not from Camiguin. They no longer had finger markings and the master list available to the enumerators were only for the province. With no other means of verification, they were recorded as unvaccinated. These findings emphasize that high quality finger markers must be provided and vaccination teams must be taught the correct way to mark the finger to prevent premature erasure.^8^ Vaccination cards should also be provided as a secondary means of validation, especially for children who are only visiting the area being surveyed. The caregiver of one unvaccinated child claimed that the vaccination team did not visit, despite the neighboring children being vaccinated. One child was unvaccinated due to the absence of a caregiver at the time of vaccination and another was due to fear from side effects. The last three reasons are related to unawareness and the need for better risk communication for the caregivers to appreciate the importance and benefits of the vaccine.

### Mobile Data Collection for LQAS

The principal investigator had no difficulty using KoBo Toolbox in creating forms and managing the data. The enumerators easily learned to use either KoBoCollect for Android or Enketo for iOS, despite being first-time users. Programming skip patterns into the form and eliminating the manual encoding of data prevented errors in recording. The Global Positioning System (GPS) feature allowed recording of the exact location of each household, which ensured that enumerators follow the randomization protocol and provided Geographic Information System (GIS) maps to show local health workers where the missed children are located. In future implementations, GIS mapping allows more efficient mop-up operations. The tool allows immediate data validation and analysis which ensures timely reports to program managers and other decision makers.

### Conclusions

The province of Camiguin has accomplished high vaccination coverage for the 1^st^ Round of synchronized Polio vaccination. LQAS is an essential methodology for immediate post-campaign assessment. Due to the randomized sampling, a generalizable conclusion can be made of vaccination coverage. It can be used to validate areas when there are concerns on the set target population and administrative coverage. Add-on questions can provide additional insight on campaign awareness and reasons for non-vaccination without significantly lengthening the duration of the survey. KoBo Toolbox can be adapted for future field data collection requirements as it is more efficient over paper-based forms.

## Data Availability

Raw data may be made available by the author upon request.

## ANNEX A

### ENUMERATION FORM

Municipality*________________

Barangay* ________________

Purok ________________

Household number ________________

Age in months during the mOPV2 campaign*

________________

*Round 1 – November 25 to December 13, 2019*

Sex*

- Male
- Female

mOPV2 vaccination status*

- Vaccinated - finger marker
- Vaccinated - vaccination card
- Vaccinated - master list
- Unvaccinated

If unvaccinated, please provide the reason

*Read all the options to the caregiver and select all that applies*

- Child was absent/away from home
- Unaware of the campaign
- Vaccine team did not visit
- Child was from a different area
- Fear of harm from the vaccine
- Child is already vaccinated
- Child was sick
- Do not know/declined to respond
- Religious beliefs
- Parent/guardian was not present at the time
- Outright refusal
- Other

Vaccination campaign awareness*

*Was the caregiver aware of the polio campaign prior to the arrival of vaccination teams?*

- Yes
- No

If aware, select the sources of information regarding the campaign

*Read all the options to the caregiver and select all that applies*

- Radio
- TV
- Streamer/Tarpaulin
- Health Workers
- Flyers
- Social Media (Facebook, Twitter, etc)
- Barangay Officials
- Relatives/Neighbors
- Other

Get coordinates*

*Please ensure that your GPS is on*

latitude (x.y °) ________________

longitude (x.y °) ________________

altitude (m) ________________

accuracy (m) ________________

Comments

*Enter other pertinent information not captured in the other questions here*

________________________________________________________________

*Required questions

